# Continued Emergence and Evolution of Omicron in South Africa: New BA.4 and BA.5 lineages

**DOI:** 10.1101/2022.05.01.22274406

**Authors:** Houriiyah Tegally, Monika Moir, Josie Everatt, Marta Giovanetti, Cathrine Scheepers, Eduan Wilkinson, Kathleen Subramoney, Sikhulile Moyo, Daniel G. Amoako, Cheryl Baxter, Christian L. Althaus, Ugochukwu J. Anyaneji, Dikeledi Kekana, Raquel Viana, Jennifer Giandhari, Richard J. Lessells, Tongai Maponga, Dorcas Maruapula, Wonderful Choga, Mogomotsi Matshaba, Simnikiwe Mayaphi, Nokuzola Mbhele, Mpaphi B. Mbulawa, Nokukhanya Msomi, NGS-SA consortium, Yeshnee Naidoo, Sureshnee Pillay, Tomasz Janusz Sanko, James E. San, Lesley Scott, Lavanya Singh, Nonkululeko A. Magini, Pamela Smith-Lawrence, Wendy Stevens, Graeme Dor, Derek Tshiabuila, Nicole Wolter, Wolfgang Preiser, Florette K. Treurnicht, Marietjie Venter, Michaela Davids, Georginah Chiloane, Adriano Mendes, Caitlyn McIntyre, Aine O’Toole, Christopher Ruis, Thomas P. Peacock, Cornelius Roemer, Carolyn Williamson, Oliver G. Pybus, Jinal Bhiman, Allison Glass, Darren P. Martin, Andrew Rambaut, Simani Gaseitsiwe, Anne von Gottberg, Tulio de Oliveira

## Abstract

South Africa’s fourth COVID-19 wave was driven predominantly by three lineages (BA.1, BA.2 and BA.3) of the SARS-CoV-2 Omicron variant of concern. We have now identified two new lineages, BA.4 and BA.5. The spike proteins of BA.4 and BA.5 are identical, and comparable to BA.2 except for the addition of 69-70del, L452R, F486V and the wild type amino acid at Q493. The 69-70 deletion in spike allows these lineages to be identified by the proxy marker of S-gene target failure with the TaqPath™ COVID-19 qPCR assay. BA.4 and BA.5 have rapidly replaced BA.2, reaching more than 50% of sequenced cases in South Africa from the first week of April 2022 onwards. Using a multinomial logistic regression model, we estimate growth advantages for BA.4 and BA.5 of 0.08 (95% CI: 0.07 - 0.09) and 0.12 (95% CI: 0.09 - 0.15) per day respectively over BA.2 in South Africa.

## Main text

Within days of being discovered in South Africa and Botswana, on November 26, 2021, the Omicron variant of SARS-CoV-2 was designated as a variant of concern (VOC) by the World Health Organization^1^. Initially, Omicron was comprised of three sister lineages, BA.1, BA.2 and BA.3. BA.1 caused most of the infections in South Africa’s fourth epidemic wave. However, as that wave receded in mid-January 2022, BA.2 became the dominant South African lineage. Despite being associated with a modest prolongation of the fourth wave, the displacement of BA.1 by BA.2 in South Africa was not associated with a significant resurgence in cases, hospital admissions or deaths. This pattern was not consistent worldwide, however, and in some countries BA.2 was responsible for a greater share of cases, hospitalizations and deaths in the Omicron wave^2–7^.

We recently identified two new Omicron lineages that have been designated BA.4 and BA.5 by the Pango Network and pango-designation v1.3 (Fig. 1A)^8,9^. Bayesian phylogenetic methods revealed that BA.4 and BA.5 are distinct from the other Omicron lineages. BA.4 and BA.5 are estimated to have originated in mid-December 2021 (95% highest posterior density [HPD] 25 November 2021 to 01 January 2022) and early January 2022 (HPD 10 December 2021 to 6 February 2022) respectively (Fig. 1A). The most recent common ancestor of BA.4 and BA.5 is estimated to have originated in mid-November 2021 (HPD 29 September 2021 to 6 December 2021) (Fig. 1A), coinciding with the emergence of the other lineages, for example BA.2 in early November 2021 (HPD: 9 October 2021 to 29 November 2021). Phylogeographic analysis suggests early dispersal of BA.4 from Limpopo to Gauteng, with later spread to other provinces (Fig. 1B); and early dispersal of BA.5 from Gauteng to KwaZulu-Natal, with more limited onward spread to other provinces (Fig. 1C).

**Figure 1:**
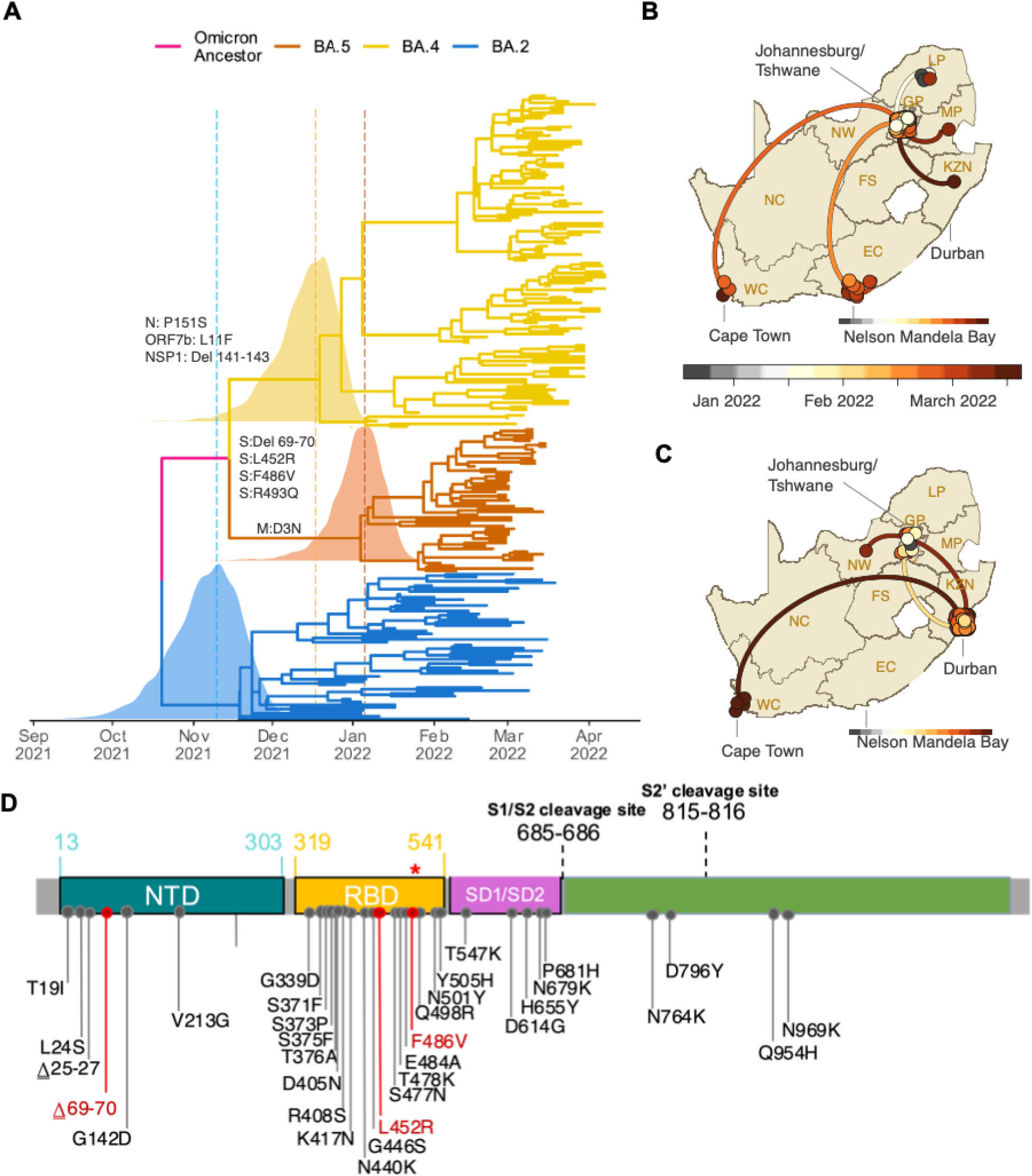
A) Time-resolved maximum clade credibility phylogeny of the BA.2, BA.4 and BA.5 lineages (n = 221, sampled between 29 December 2021 and 7 April 2022). Mutations that characterize the lineages are indicated on the branch at which each first emerged. The posterior distribution of the time of the most recent common ancestor (TMRCA) is also shown for BA.2, BA.4 and BA.5. B) Spatiotemporal reconstruction of the spread of the BA.4 lineage in South Africa. C) Spatiotemporal reconstruction of the spread of the BA.5 lineage in South Africa. In B and C, circles represent nodes of the maximum clade credibility phylogeny, coloured according to their inferred time of occurrence (scale shown). Solid curved lines denote the links between nodes and the directionality of movement (anti-clockwise along the curve). D) Amino acid mutations in the spike gene of the BA.4 and BA.5 lineages. Mutations that differ from BA.2 are denoted in red, including the wild-type amino acid at position Q493 (denoted by the red *).

BA.4 and BA.5 have identical spike proteins, most comparable to BA.2. Relative to BA.2, BA.4 and BA.5 have the additional spike mutations 69-70del, L452R, F486V and wild type amino acid at position Q493 (Fig 1D). Outside of spike, BA.4 has the additional mutations at ORF7b:L11F and N:P151S and a triple amino acid deletion in NSP1:141-143del whilst BA.5 has the M:D3N mutation. Relative to BA.2, BA.5 has additional reversions at ORF6:D61 and nucleotide positions 26858 and 27259. In addition, BA.4 and BA.5 have a nuc:G12160A synonymous mutation in NSP8 that was present in Epsilon (B.1.429) and has arisen in BA.2 in some locations (Extended Data Fig. 1). BA.4 and BA.5 have identical mutational patterns in the 5’ genome region (from ORF1ab to Envelope) yet exhibit genetic divergence in the 3’ region (from M to the 3’ genome end). This suggests that BA.4 and BA.5 may be related by a recombination event, with breakpoint between the E and M genes, prior to their emergence into the general population. This scenario is somewhat similar to the relationship between BA.3 and BA.1/BA.2 which also exhibit apparent ancestral recombination. Using the RASCL pipeline ^10^ we found no compelling evidence of natural selection acting on the S-genes of viruses in either the BA.4 or BA.5 lineages.

It is currently unknown how differences in the mutation profiles of BA.4 and BA.5, relative to BA.2, will impact on the phenotype. Changes at spike amino acids 452, 486 and 493 are likely to influence human angiotensin-converting enzyme-2 (hACE2) and antibody binding. The 452 residue is in immediate proximity to the interaction interface of the hACE2 receptor. The L452R mutation has been associated with an increased affinity for receptor binding with a resultant increased infectivity^11,12^. The L452R mutation is also present in the Delta, Kappa and Epsilon variants (and L452Q in Lambda), and mutations at this position have been associated with a reduction in neutralization by monoclonal antibodies (particularly class 2 antibodies) and polyclonal sera^13–15^. Mutations at this position (L452R/M/Q) have also arisen independently in at least four BA.2 sublineages in different parts of the world, most notably BA.2.12.1 (L452Q) which has become dominant in New York State.

Before the emergence of BA.4and BA.5, F486V had been observed only in 54 of 10 million publicly available genome sequences in GISAID (https://cov-spectrum.org/explore/World/AllSamples/AllTimes/variants?aaMutations=S%3AF486V&). Selection analyses focusing on ratios of non-synonymous and synonymous substitution rates at individual codons have indicated that, since December 2020 S:486 has been evolving under strong negative selection favouring the F state at this site (i.e., the amino acid that is found in Wuhan-Hu-1) (Extended Data Fig. 2). Although rare, the F486L mutation has been observed in some viral lineages found infecting minks and in human cases linked to mink farms and has been shown to directly enhance entry into cells expressing mink/ferret ACE2 ^16^. When binding to hACE2, spike amino acid F486 interacts with hACE2 residues L79, M82, and Y83, which collectively comprise a hotspot for ACE2 differences between mammalian species^17,18^. Mutations at F486 are associated with a reduction in neutralising activity by class 1 (and some class 2) neutralising antibodies and by polyclonal sera^13–15^. Deep mutational scanning suggests that F486 is a key site for escape of vaccine- and infection-elicited RBD-targeted antibodies, including those still able to neutralize Omicron/BA.1 (https://jbloomlab.github.io/SARS2_RBD_Ab_escape_maps/escape-calc/)^19^.

**Figure 2.**
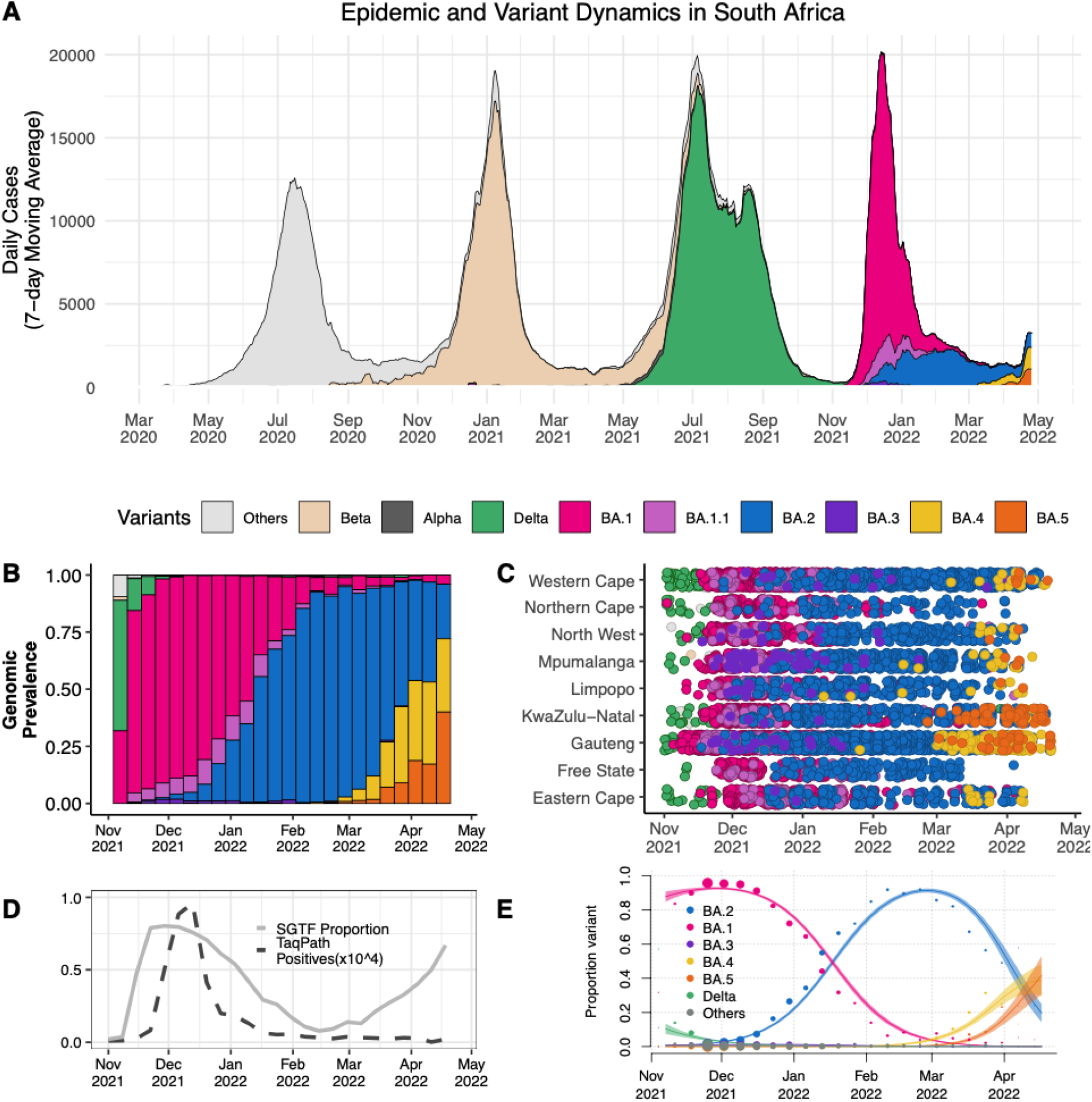
A) The progression of the 7-day rolling average of daily reported case numbers in South Africa over two years of the epidemic (April 2020 – April 2022). Daily cases are coloured by the inferred proportion of SARS-CoV-2 variants prevalent at a particular period in the epidemic. B) Changes in the genomic prevalence of Omicron lineages in South Africa from November 2021 (when BA.1 dominated) to April 2022 (when BA.4 and BA.5 were increasing in frequency). C) The count of Omicron lineage genomes per province of South Africa over November 2021 – April 2022. BA.4 and BA.5 have been detected in seven of the nine provinces.D) Changes in the proportion of positive TaqPath qPCR tests exhibiting SGTF from November 2021 to April 2022. The number of TaqPath positives are to the order of 10^^4^ to the scale shown.E) Modelled linear proportions of the Omicron lineages in South Africa. BA.1 rapidly outcompeted Delta in November 2021 and was then superseded by BA.2 in early 2022. BA.4 and BA.5 appear to be swiftly replacing BA.2 in South Africa. Model fits are based on a multinomial logistic regression and dot size represents the weekly sample size.

The S:69-70del means BA.4 and BA.5 can again be presumptively identified (against a background of BA.2 infection) using the proxy marker of S-gene target failure (SGTF) with the TaqPath™ COVID-19 qPCR assay (Thermo Fisher Scientific, Waltham, MA, USA). SGTF was successfully used to track the early spread of BA.1 (which also demonstrates SGTF), later also enabling discrimination between BA.1 and BA.2 infections, since BA.2 viruses generally lack the S:69-70del^20^. Recent data from public laboratories in South Africa suggest that the proportion of positive PCR tests with SGTF has been increasing since early March, suggesting that BA.4 and BA.5 may be responsible for a growing share of recently confirmed cases (Fig. 2E). To assess the validity of SGTF for identifying BA.4/BA.5, we performed qPCR with the TaqPath™ assay on 296 unselected samples submitted for sequencing to KwaZulu-Natal Research Innovation and Sequencing Platform (KRISP) from Gauteng, Eastern Cape and KwaZulu-Natal collected between 6 January and 3 April 2022. Of the 296 samples processed, we had a paired valid qPCR result and sequence for 198. Of the 77 samples with SGTF on qPCR, 66 were BA.4 or BA.5, nine were BA.1, and two were BA.2. No BA.4 and BA.5 genomes were S-gene target positive on qPCR (Extended Data Table 1). These results suggest that SGTF surveillance (where the assay is available) may for now be a reasonable proxy to identify BA.4 and BA.5 for countries with a low prevalence of BA.1.

At the time of writing, we have confirmed BA.4 and/or BA.5 in seven provinces in South Africa (Eastern Cape, Gauteng, KwaZulu-Natal, Limpopo, Mpumalanga, North West and Western Cape) in samples collected between 1 January 2022 and 20 April 2022 (Fig. 2B). In the two most populous provinces in South Africa, Gauteng and KwaZulu-Natal, BA.4 and BA.5 have rapidly replaced BA.2, and are responsible for approximately 60-75% of sequenced cases by the week starting 18 April 2022 (Extended Data Fig. 3). These estimates are based on unselected sampling for genomic surveillance (samples not selected based on SGTF or genotyping). The data suggest geographic heterogeneity in the distribution of these two new lineages, with growth predominantly of BA.4 in Gauteng and BA.5 in KwaZulu-Natal (Extended Data Fig. 3). Internationally, by 20 April 2022, BA.4 had also been detected in a small number of samples in neighbouring Botswana (estimated prevalence <5%), in Europe (Belgium, Denmark, France, Switzerland and UK), and USA, while BA.5 has also been detected in Europe (Austria, France, Germany, Portugal and UK), USA and Hong Kong.

We estimated that Omicron BA.4 and BA.5 had a daily growth advantage of 0.08 (95% confidence interval [CI] 0.07 - 0.09) and 0.12 (95% CI 0.09 - 0.15) respectively, relative to BA.2 in South Africa in April 2022 (Fig. 2F). These estimates are similar to the estimated daily growth advantage of 0.07 (95% CI: 0.07 – 0.08) of BA.2 over BA.1 in February 2022 (Fig 2F). The growth advantage of Omicron BA.4 and BA.5 could be mediated by either (i) an increase in its intrinsic transmissibility relative to other variants, (ii) an increase relative to other variants in its capacity to infect, and be transmitted from, previously infected and vaccinated individuals or (iii) both. The estimated time to most recent common ancestor for both BA.4 and BA.5 (mid-November 2021) argues against the first option because that suggests both lineages would have been circulating throughout the period dominated by BA.1 and then BA.2 without exhibiting a transmission advantage. The observation that both BA.4 and BA.5 (and many lineages within them) have recently started to grow in frequency suggests the growth advantage is recent and uniform across these lineages. It is estimated that almost all of the South African population has some degree of immunity to SARS-CoV-2, provided by a complex mixture of vaccination and prior infections with wild-type, Beta, Delta, and Omicron (particularly BA.1) (Fig 2A)^21,22^. Given that the transmission advantage becomes apparent approximately four months from the start of the Omicron wave, it is plausible that waning immunity (particularly that acquired from BA.1 infection) is an important contributory factor. This would also suggest that the effects of these different Omicron lineages may differ by location, depending on the immune landscape and particularly the patterns of exposure to BA.1 and BA.2.

It remains unclear how large an effect this shift in the distribution of different Omicron lineages will have on the epidemic in South Africa and elsewhere in the world. Although the incidence of reported cases (Fig. 2A) and the proportions of positive qPCR and antigen tests were relatively low (5-10%) through to early April 2022, these indicators began to rise from mid-April 2022, and at the time of writing there are early signs of rising hospital admissions in some provinces. Work is underway to characterise disease severity and immune escape.

There remains some uncertainty about the origin of the different Omicron lineages and phylogenetic inference is limited by the relatively low sampling coverage in our genomic surveillance. Whilst the Bayesian phylogenetic methods employed here suggest that BA.4 and BA.5 are independent lineages that originated around the same time as BA.1-BA.3, other methods suggest they could have descended from BA.2. Further sequencing (particularly samples from Gauteng and neighbouring provinces) may help to provide more clarity. Nevertheless, the continued discovery of genetically diverse Omicron lineages shifts the level of support for hypotheses regarding their origin, from an unsampled location to a discrete reservoir, such as human chronic infections (or even a network of chronic human infections) and/or animal reservoirs, potentially contributing to further evolution and dispersal of the virus. We are actively investigating the potential of a yet unidentified animal reservoir in the region. To date, the only reverse zoonoses cases reported from the African region were in African lions and a puma in a private zoo in Johannesburg, South Africa ^23^. Although these are unlikely species to play a role in the emergence of new variants, it is a reminder of the susceptibility of certain wildlife species to infections from humans. Following the emergence of Omicron, the World Organisation for Animal Health released a statement calling for enhanced surveillance in animals to identify the origin of new variants ^24^.

In conclusion, we have identified two new Omicron lineages (BA.4 and BA.5), which seem to be associated with a resurgence in infections in South Africa approximately four months on from the start of the Omicron wave. This once again highlights the importance of continued global genomic surveillance and variant analysis in real-time to characterize the continuing evolution of SARS-CoV-2.

## Data Availability

All data produced are available online at GISAID database.

## Online Methods

### Epidemiological dynamics

We analysed daily cases of SARS-CoV-2 in South Africa up to 25 April 2022 from publicly released data provided by the National Department of Health and the National Institute for Communicable Diseases. This was accessible through the repository of the Data Science for Social Impact Research Group at the University of Pretoria (https://github.com/dsfsi/covid19za)^25,26^. The National Department of Health releases daily updates on the number of confirmed new cases, deaths and recoveries, with a breakdown by province.

### Sampling of SARS-CoV-2

As part of the NGS-SA^27^, seven sequencing hubs receive randomly selected samples for sequencing every week according to approved protocols at each site. These samples include remnant nucleic acid extracts or remnant nasopharyngeal and oropharyngeal swab samples from routine diagnostic SARS-CoV-2 PCR testing from public and private laboratories in South Africa. We analysed SARS-CoV-2 genomes generated from samples collected between 1 November 2021 and 20th April 2022.

### Ethical statement

The project was approved by University of KwaZulu–Natal Biomedical Research Ethics Committee (ref. BREC/00001510/2020), the University of the Witwatersrand Human Research Ethics Committee (HREC) (ref. M180832), Stellenbosch University HREC (ref. N20/04/008_COVID-19) and the University of Cape Town HREC (ref. 383/2020). Individual participant consent was not required for the genomic surveillance. This requirement was waived by the Research Ethics Committees.

### Whole-genome sequencing and genome assembly

RNA was extracted on an automated Chemagic 360 instrument, using the CMG-1049 kit (Perkin Elmer). The RNA was stored at −80□°C before use. Libraries for whole-genome sequencing were prepared using either the Oxford Nanopore Midnight protocol with Rapid Barcoding or the Illumina COVIDseq Assay.

### Illumina Miseq/NextSeq

For the Illumina COVIDseq assay, the libraries were prepared according to the manufacturer’s protocol. In brief, amplicons were tagmented, followed by indexing using the Nextera UD Indexes Set A. Sequencing libraries were pooled, normalized to 4□nM and denatured with 0.2□N sodium acetate. A 8□pM sample library was spiked with 1% PhiX (PhiX Control v3 adaptor-ligated library used as a control). We sequenced libraries using the 500-cycle v2 MiSeq Reagent Kit on the Illumina MiSeq instrument (Illumina). On the Illumina NextSeq 550 instrument, sequencing was performed using the Illumina COVIDSeq protocol (Illumina), an amplicon-based next-generation sequencing approach. The first-strand synthesis was performed using random hexamers primers from Illumina and the synthesized cDNA underwent two separate multiplex PCR reactions. The pooled PCR amplified products were processed for tagmentation and adapter ligation using IDT for Illumina Nextera UD Indexes. Further enrichment and clean-up was performed according to protocols provided by the manufacturer (Illumina). Pooled samples were quantified using the Qubit 3.0 or 4.0 fluorometer (Invitrogen) and the Qubit dsDNA High Sensitivity assay kit according to the manufacturer’s instructions. The fragment sizes were analysed using the TapeStation 4200 (Invitrogen). The pooled libraries were further normalized to 4□nM concentration, and 25□μl of each normalized pool containing unique index adapter sets was combined into a new tube. The final library pool was denatured and neutralized with 0.2□N sodium hydroxide and 200□mM Tris-HCl (pH□7), respectively. Sample library (1.5□pM) was spiked with 2% PhiX. Libraries were loaded onto a 300-cycle NextSeq 500/550 HighOutput Kit v2 and run on the Illumina NextSeq 550 instrument (Illumina).

### Midnight protocol

For Oxford Nanopore sequencing, the Midnight primer kit was used as described previously54. cDNA synthesis was performed on the extracted RNA using the LunaScript RT mastermix (New England BioLabs) followed by gene-specific multiplex PCR using the Midnight primer pools, which produce 1,200□bp amplicons that overlap to cover the 30□kb SARS-CoV-2 genome. Amplicons from each pool were pooled and used neat for barcoding with the Oxford Nanopore Rapid Barcoding kit according to the manufacturer’s protocol. Barcoded samples were pooled and bead-purified. After the bead clean-up, the library was loaded on a prepared R9.4.1 flow-cell. A GridION X5 or MinION sequencing run was initiated using MinKNOW software with the base-call setting switched off.

### Ion Torrent Genexus Integrated Sequencer methodology for rapid whole-genome sequencing of SARS-CoV-2

Viral RNA was extracted using the MagNA Pure 96 DNA and Viral Nucleic Acid kit on the automated MagNA Pure 96 system (Roche Diagnostics) according to the manufacturer’s instructions. Extracts were then screened by quantitative PCR to acquire the mean cycle threshold (Ct) values for the SARS-CoV-2 N and ORF1ab genes using the TaqMan 2019-nCoV assay kit v1 (Thermo Fisher Scientific) on the ViiA7 Real-time PCR system (Thermo Fisher Scientific) according to the manufacturer’s instructions. Extracts were sorted into batches of n□=□8 within a Ct range difference of 5 for a maximum of two batches per run. Extracts with <200 copies were sequenced using the low viral titre protocol. Next-generation sequencing was performed using the Ion AmpliSeq SARS-CoV-2 Research Panel on the Ion Torrent Genexus Integrated Sequencer (Thermo Fisher Scientific), which combines automated cDNA synthesis, library preparation, templating preparation and sequencing within 24□h. The Ion Ampliseq SARS-CoV-2 Research Panel consists of two primer pools targeting 237 amplicons tiled across the SARS-CoV-2 genome providing >99% coverage of the SARS-CoV-2 genome (∼30□kb) and an additional five primer pairs targeting human expression controls. The SARS-CoV-2 amplicons range from 125□bp to 275□bp in length. TRINITY was used for de novo assembly and the Iterative Refinement Meta-Assembler (IRMA) was used for genome assisted assembly as well as FastQC for quality checks.

### Genome assembly

We assembled paired-end and Nanopore .fastq reads using Genome Detective v.1.132 (https://www.genomedetective.com), which was updated for the accurate assembly and variant calling of tiled primer amplicon Illumina or Oxford Nanopore reads, and the Coronavirus Typing Tool55. For Illumina assembly, the GATK HaploTypeCaller --min-pruning 0 argument was added to increase mutation calling sensitivity near sequencing gaps. For Nanopore, low-coverage regions with poor alignment quality (<85% variant homogeneity) near sequencing/amplicon ends were masked to be robust against primer drop-out experienced in the spike gene, and the sensitivity for detecting short inserts using a region-local global alignment of reads was increased. We also used the wf_artic (ARTIC SARS-CoV-2) pipeline as built using the Nextflow workflow framework56. In some instances, mutations were confirmed visually with .bam files using Geneious v.2020.1.2 (Biomatters). The reference genome used throughout the assembly process was NC_045512.2 (numbering equivalent to MN908947.3).

Raw reads from the Illumina COVIDSeq protocol were assembled using the Exatype NGS SARS-CoV-2 pipeline v.1.6.1 (https://sars-cov-2.exatype.com/). This pipeline performs quality control on reads and then maps the reads to a reference using Examap. The reference genome used throughout the assembly process was NC_045512.2 (accession number: MN908947.3).

Several of the initial Ion Torrent genomes contained a number of frameshifts, which caused unknown variant calls. Manual inspection revealed that these were probably sequencing errors resulting in mis-assembled regions (probably due to the known error profile of Ion Torrent sequencers)57. To resolve this, the raw reads from the IonTorrent platform were assembled using the SARSCoV2 RECoVERY (Reconstruction of Coronavirus Genomes & Rapid Analysis) pipeline implemented in the Galaxy instance ARIES (https://aries.iss.it). This pipeline fixed the observed frameshifts, confirming that they were artefacts of mis-assembly; this subsequently resolved the variant calls. The Exatype and RECoVERY pipelines each produce a consensus sequence for each sample. These consensus sequences were manually inspected and polished using Aliview v.1.27 (http://ormbunkar.se/aliview/).

All of the sequences passing internal quality control were deposited in GISAID (https://www.gisaid.org/), and the GISAID accession identifiers are included as part of Extended Data Table 1.

### Phylogenetic analysis

We initially analysed genomes from South Africa against the global reference dataset using a custom pipeline based on a local version of NextStrain (https://github.com/nextstrain/ncov)^28^. The pipeline contains several Python scripts that manage the analysis workflow. It performs an alignment of genomes in NextAlign ^29^, phylogenetic tree inference in IQ-Tree V1.6.9^30^, tree dating and ancestral state construction and annotation (https://github.com/nextstrain/ncov).

The initial phylogenetic analysis enabled us to identify clusters corresponding to the BA.4 (n=120) and BA.5 (n=51) lineages. We extracted these clusters and constructed a preliminary maximum-likelihood tree with a subset of BA.2 sequences (n=52) in IQ-tree. We inspected this maximum-likelihood tree in TempEst v.1.5.3^31^ for the presence of a temporal or molecular clock signal. Linear regression of root-to-tip genetic distances against sampling dates indicated that the SARS-CoV-2 sequences evolved in a relatively strong clock-like manner (correlation coefficient = 0.6, R^2^ = 0.4) (Extended Data Fig. 4).

We then estimated time-calibrated phylogenies using the Bayesian software package BEAST v.1.10.4^32^. For this analysis, we used the strict molecular clock model, the HKY+I+G, nucleotide substitution model and the exponential growth coalescent model^33^. We computed Markov chain Monte Carlo (MCMC) in duplicate runs of 20 million states each, sampling every 2,000 steps. Convergence of MCMC chains was checked using Tracer v.1.7.1^34^. Maximum clade credibility trees were summarized from the MCMC samples using TreeAnnotator after discarding 10% as burn-in. The phylogenetic trees were visualized using ggplot and ggtree^35,36^.

### Phylogeographic analysis

To model phylogenetic diffusion of the new cluster across the country, we used a flexible relaxed random walk diffusion model that accommodates branch-specific variation in rates of dispersal with a Cauchy distribution^37^. For each sequence, latitude and longitude were attributed to the most precise district or provincial information available and linked to the diagnostic sample.

As described in ‘Phylogenetic analysis’, MCMC chains were run in duplicate for 10 million generations and sampled every 1,000 steps, with convergence assessed using Tracer v.1.7.1. Maximum clade credibility trees were summarized using TreeAnnotator after discarding 10% as burn-in. We used the R package seraphim^38^ to extract and map spatiotemporal information embedded in posterior trees.

### Lineage classification

We used a previously proposed dynamic lineage classification method^39^ from the ‘Phylogenetic Assignment of Named Global Outbreak Lineages’ (pangolin) software suite v4.0.6 with the -- Usher option (https://github.com/cov-lineages/pangolin) ^40^. This is aimed at identifying the most epidemiologically important lineages of SARS-CoV-2 at the time of analysis, enabling researchers to monitor the epidemic in a particular geographic region. A lineage is a linear chain of viruses in a phylogenetic tree showing connection from the ancestor to the last descendant. Variant refers to a genetically distinct virus with different mutations to other viruses.

### Selection analysis

To identify which (if any) of the observed mutations in the spike protein was most likely to increase viral fitness, we used the natural selection analysis of SARS-CoV-2 pipeline (https://observablehq.com/@spond/revised-sars-cov-2-analytics-page). This pipeline examines the entire global SARS-CoV-2 nucleotide sequence dataset for evidence of: (i) polymorphisms having arisen in multiple epidemiologically unlinked lineages that have statistical support for non-neutral evolution (mixed effects model of evolution)^41^, (ii) sites at which these polymorphisms have support for a greater-than-expected ratio of nonsynonymous-to-synonymous nucleotide substitution rates on internal branches of the phylogenetic tree (fixed-effects likelihood)^42^ and (iii) whether these polymorphisms have increased in frequency in the regions of the world in which they have occurred.

### Estimating transmission advantage

We analysed 12,528 SARS-CoV-2 sequences from South Africa generated in this study and uploaded to GISAID with sample collection dates from 1 November 2021 to 20 April 2022 ^43^. We used a multinomial logistic regression model to estimate the growth advantage of Omicron BA.2 lineage compared with BA.1, BA.4 and BA.5 lineages at the time point at which the proportion of Omicron BA.4 and BA.5 collectively reached 50% ^44,45^. We fitted the model using the multinom function of the nnet package and estimated the growth advantage using the package emmeans in R ^46^.

### S-Gene Target Failure Monitoring

SGTF monitoring is performed through analysing SARS-CoV-2 laboratory test results from nasopharyngeal specimens received from the public health sector and referred for PCR testing undertaken by the National Health Laboratory Service (NHLS) in South Africa. The NHLS has a single laboratory information system connecting laboratory testing platforms to a corporate data warehouse, where data can be mined in near real-time. The TaqPath™ COVID-19 [Thermo Fisher Scientific, Waltham, MA, USA] assay accounts for around 20% of NHLS PCR tests performed, with around half of those performed in Gauteng. The TaqPath assay targets three gene regions, ORF1ab, N and S, with the lack of probe fluorescence of the latter culminating in S-gene target failure (SGTF). In Fig 2D, we analysed and plotted the weekly number of TaqPath positive tests as well as the proportion of the positive tests with SGTF (defined as samples with non-detectable S gene target and either N or ORF1ab gene positive with CT value <30.

### Validation of S-Gene Target status as proxy for BA.4 and BA.5

Using a subset of unselected samples submitted to the KRISP sequencing laboratory, we compared the S-gene target status to the genome lineage assignment. Briefly, RNA was extracted from nasopharyngeal swabs in viral transport media using the CMG-1033-S kit (Chemagen, PerkinElmer, Baesweiler, Germany). 10µl of purified RNA was then amplified using the TaqPath COVID-19 CE-IVD RT-PCR kit (ThermoFisher Scientific, Waltham, MA, USA) and analysed on the Design & Analysis software v2.4. SGTF was denoted by lack of amplification of the S-gene target, with successful amplification of both the remaining ORF1ab and N-gene targets (Ct ≤ 30).

## NGS-SA consortium author list

Phillip A. Bester, Maciej F. Boni, Mohammed Chand, Kutlo Macheke, Rachel Colquhoun, Michaela Davids, Koen Deforche, Deelan Doolabh, Louis du Plessis, Susan Engelbrecht, Diana Hardie, Verity Hill, Nei-Yuan Hsiao, Arash Iranzadeh, Arshad Ismail, Charity Joseph, Rageema Joseph, Legodile Koopile, Faith Hungwe, Nokuthula Ndlovu, Lesego Kuate-Lere, Oluwakemi Laguda-Akingba, Onalethatha Lesetedi-Mafoko, Shahin Lockman, Nkhensani Mtileni, Ashlyn S.C. Davis Makama, Annabel Enoch, Luicer Olubayo, Arisha Maharaj, Boitshoko Mahlangu, Kamela Mahlakwane, Gert van Zyl, Mathilda Claassen, Shannon Wilson, Zinhle Makatini, Gert Marais, Koleka Mlisana, Anele Mnguni, Thabo Mohale, Kgomotso Moruisi, Mosepele Mosepele, Kereng V. Masupu, Gerald Motsatsi, Modisa S. Motswaledi, Thongbotho Mphoyakgosi, Noxolo Ntuli, Martin Nyaga, Lucier Olubayo, Botshelo Radibe, Yajna Ramphal, Upasana Ramphal, Roger Shapiro, Naume Tebeila, Wilhelmina Strasheim, Joseph Tsui, Stephanie van Wyk, Steven Weaver, Nicole Wolter, Alexander E. Zarebski, Boitumelo Zuze, Dominique Goedhals, Armand (Phillip) Bester, Martin Nyaga and Peter Mwangi.

## Funding Information

This research was supported by the South African Medical Research Council (SAMRC) with funds received from the National Department of Health. Sequencing activities for NICD are supported by a conditional grant from the South African National Department of Health as part of the emergency COVID-19 response; a cooperative agreement between the National Institute for Communicable Diseases of the National Health Laboratory Service and the United States Centers for Disease Control and Prevention (CDC)(grant number 5 U01IP001048-05-00; 1 NU51IP000930-01-00); the African Society of Laboratory Medicine (ASLM) and Africa Centers for Disease Control and Prevention through a sub-award from the Bill and Melinda Gates Foundation grant number INV-018978; the UK Foreign, Commonwealth and Development Office and Wellcome (Grant no 221003/Z/20/Z); and the UK Department of Health and Social Care and managed by the Fleming Fund and performed under the auspices of the SEQAFRICA project. This research was also supported by The Coronavirus Aid, Relief, and Economic Security Act (CARES ACT) through the CDC and the COVID International Task Force (ITF) funds through the CDC under the terms of a subcontract with the African Field Epidemiology Network (AFENET) AF-NICD-001/2021. Sequencing activities at KRISP and CERI are supported in part by grants from the World Health Organization, the Abbott Pandemic Defense Coalition (APDC), the National Institute of Health USA (U01 AI151698) for the United World Antivirus Research Network (UWARN) and the INFORM Africa project through IHVN (U54 TW012041), and the South African Department of Science and Innovation (SA DSI) and the SAMRC under the BRICS JAF #2020/049.

## Acknowledgements

We thank additional members from originating and sequencing laboratories in South Africa, listed as part of the NGS-SA consortium authors, that helped to generate and made public the SARS-CoV-2 sequences (through GISAID) used as reference dataset in this study (a complete list of individual contributors of sequences is provided in Supplementary Table S1).

## Data Availability Statement

All of the SARS-CoV-2 genomes generated and presented in this manuscript are publicly accessible through the GISAID platform (https://www.gisaid.org/). The GISAID accession identifiers of the sequences analysed in this study are provided as part of Supplementary Table S1. Other raw data for this study are provided as a supplementary dataset at https://github.com/krisp-kwazulu-natal/SARSCoV2_South_Africa_Omicron_BA4_BA5. The reference SARS-CoV-2 genome (MN908947.3) was downloaded from the NCBI database (https://www.ncbi.nlm.nih.gov/).

## Code Availability Statement

All custom scripts to reproduce the analyses and figures presented in this Article are available at https://github.com/krisp-kwazulu-natal/SARSCoV2_South_Africa_Omicron_BA4_BA5.

## Author Contributions

Genomic or Diagnostics data generation: H.T., M.Moir, J.E., C.S., K.S., S.Moyo, D.G.A., U.J.A., D.K., R.V., J.G., T.M., D.M., W.C., M.Matshaba, S.Mayaphi, N.Mbhele, N.B.M., Y.N., S.P., T.J.S., J.E.S, L.Scott, L.Singh, N.A.M., P.S.L., W.S., G.D., D.T., N.W., W.P., F.K.T, C.W., J.B.

Sample collection and metadata curation: N.Msomi, M.V., K.S., F.K.T., M.D., G.C., A.M., C.M.,

Data analysis: H.T., M.Moir, M.G., E.W., J.E., D.G.A., K.S., A.OT., C.R., T.P.P., C.R., O.G.P, D.P.M., A.R.

Study design and data interpretation: H.T., M.Moir, E.W., C.L.A, R.J.L., C.W., O.G.P, J.B., A.G., D.P.M., A.R., S.G., A.vG., T.dO

Manuscript writing: H.T., M.Moir, M.G., E.W., C.B., R.J.L., T.dO

All of the authors reviewed the manuscript.

## Notes

### Competing Interest Statement

The authors have declared no competing interest.

### Author Declarations

The project was approved by University of KwaZulu-Natal Biomedical Research Ethics Committee (ref. BREC/00001510/2020), the University of the Witwatersrand Human Research Ethics Committee (HREC) (ref. M180832), Stellenbosch University HREC (ref. N20/04/008 COVID-19) and the University of Cape Town HREC (ref. 383/2020). Individual participant consent was not required for the genomic surveillance. This requirement was waived by the Research Ethics Committees.

## References

1. Viana, R. et al. Rapid epidemic expansion of the SARS-CoV-2 Omicron variant in southern Africa. Nature 603, 679–686 (2022).

2. Lyngse, F. P. et al. Transmission of SARS-CoV-2 Omicron VOC subvariants BA.1 and BA.2: Evidence from Danish Households. medRxiv (2022) doi:10.1101/2022.01.28.22270044.

3. Rahimi, F. & Talebi Bezmin Abadi, A. The Omicron subvariant BA.2: Birth of a new challenge during the COVID-19 pandemic. Int. J. Surg. 99, 106261 (2022).

4. Fonager, J. et al. Molecular epidemiology of the SARS-CoV-2 variant Omicron BA.2 sub-lineage in Denmark, 29 November 2021 to 2 January 2022. Euro Surveill. 27, (2022).

5. Dai, Y. Rapid epidemic expansion of the SARS-CoV-2 Omicron BA.2 subvariant during China’s largest outbreaks. Res. Sq. (2022) doi:10.21203/rs.3.rs-1516063/v3.

6. Chen, L.-L. et al. Contribution of low population immunity to the severe Omicron BA.2 outbreak in Hong Kong. Res. Sq. (2022) doi:10.21203/rs.3.rs-1512533/v1.

7. Hirotsu, Y. et al. SARS-CoV-2 Omicron sublineage BA.2 replaces BA.1.1: genomic surveillance in Japan from September 2021 to March 2022. medRxiv (2022) doi:10.1101/2022.04.05.22273483.

8. O’Toole, Å., Pybus, O. G., Abram, M. E., Kelly, E. J. & Rambaut, A. Pango lineage designation and assignment using SARS-CoV-2 spike gene nucleotide sequences. BMC Genomics 23, 121 (2022).

9. Rambaut, A. et al. A dynamic nomenclature proposal for SARS-CoV-2 lineages to assist genomic epidemiology. Nat. Microbiol. 5, 1403–1407 (2020).

10. Lucaci, A. G. et al. RASCL: Rapid Assessment Of SARS-CoV-2 Clades Through Molecular Sequence Analysis. BioRxiv (2022) doi:10.1101/2022.01.15.476448.

11. Motozono, C. et al. SARS-CoV-2 spike L452R variant evades cellular immunity and increases infectivity. Cell Host Microbe 29, 1124-1136.e11 (2021).

12. Chen, J., Wang, R., Wang, M. & Wei, G.-W. Mutations Strengthened SARS-CoV-2 Infectivity. J. Mol. Biol. 432, 5212–5226 (2020).

13. Greaney, A. J. et al. Mapping mutations to the SARS-CoV-2 RBD that escape binding by different classes of antibodies. Nat. Commun. 12, 4196 (2021).

14. Greaney, A. J. et al. Comprehensive mapping of mutations in the SARS-CoV-2 receptor-binding domain that affect recognition by polyclonal human plasma antibodies. Cell Host Microbe 29, 463-476.e6 (2021).

15. Greaney, A. J. et al. Complete Mapping of Mutations to the SARS-CoV-2 Spike Receptor-Binding Domain that Escape Antibody Recognition. Cell Host Microbe 29, 44-57.e9 (2021).

16. Zhou, J. et al. Mutations that adapt SARS-CoV-2 to mink or ferret do not increase fitness in the human airway. Cell Rep. 38, 110344 (2022).

17. Lan, J. et al. Structure of the SARS-CoV-2 spike receptor-binding domain bound to the ACE2 receptor. Nature 581, 215–220 (2020).

18. Shang, J. et al. Structural basis of receptor recognition by SARS-CoV-2. Nature 581, 221– 224 (2020).

19. Greaney, A. J., Starr, T. N. & Bloom, J. D. An Antibody-Escape Estimator for Mutations to the SARS-CoV-2 Receptor-Binding Domain. Virus Evol. (2022) doi:10.1093/ve/veac021.

20. Scott, L. et al. Track Omicron’s spread with molecular data. Science 374, 1454–1455 (2021).

21. Sun, K. et al. Persistence of SARS-CoV-2 immunity, Omicron’s footprints, and projections of epidemic resurgences in South African population cohorts. medRxiv (2022) doi:10.1101/2022.02.11.22270854.

22. Madhi, S. A. et al. Population Immunity and Covid-19 Severity with Omicron Variant in South Africa. N. Engl. J. Med. 386, 1314–1326 (2022).

23. Koeppel, K. N. et al. SARS-CoV-2 Reverse Zoonoses to Pumas and Lions, South Africa. Viruses 14, (2022).

24. Statement from the Advisory Group on SARS-CoV-2 Evolution in Animals concerning the origins of Omicron variant - OIE - World Organisation for Animal Health. https://www.oie.int/en/document/statement-from-the-advisory-group-on-sars-cov-2-evolution-in-animals-concerning-the-origins-of-omicron-variant/.

## Supplementary References

25. Marivate, V. & Combrink, H. M. Use of Available Data To Inform The COVID-19 Outbreak in South Africa: A Case Study. Data Sci. J. 19, (2020).

26. Marivate, V. et al. Coronavirus disease (COVID-19) case data - South Africa. Zenodo (2020) doi:10.5281/zenodo.3819126.

27. Msomi, N., Mlisana, K., de Oliveira, T. & Network for Genomic Surveillance in South Africa writing group. A genomics network established to respond rapidly to public health threats in South Africa. Lancet Microbe 1, e229–e230 (2020).

28. Hadfield, J. et al. Nextstrain: real-time tracking of pathogen evolution. Bioinformatics 34, 4121–4123 (2018).

29. GitHub - neherlab/nextalign: □ Viral genome reference alignment. https://github.com/neherlab/nextalign.

30. Nguyen, L.-T., Schmidt, H. A., von Haeseler, A. & Minh, B. Q. IQ-TREE: a fast and effective stochastic algorithm for estimating maximum-likelihood phylogenies. Mol. Biol. Evol. 32, 268–274 (2015).

31. Rambaut, A., Lam, T. T., Max Carvalho, L. & Pybus, O. G. Exploring the temporal structure of heterochronous sequences using TempEst (formerly Path-O-Gen). Virus Evol. 2, vew007 (2016).

32. Suchard, M. A. et al. Bayesian phylogenetic and phylodynamic data integration using BEAST 1.10. Virus Evol. 4, vey016 (2018).

33. Griffiths, R. C. & Tavaré, S. Sampling theory for neutral alleles in a varying environment. Philos. Trans. R. Soc. Lond. B Biol. Sci. 344, 403–410 (1994).

34. Rambaut, A., Drummond, A. J., Xie, D., Baele, G. & Suchard, M. A. Posterior summarization in bayesian phylogenetics using tracer 1.7. Syst. Biol. 67, 901–904 (2018).

35. Wickham, H. ggplot2. WIREs Comp Stat 3, 180–185 (2011).

36. Yu, G. Using ggtree to Visualize Data on Tree-Like Structures. Curr. Protoc. Bioinformatics 69, e96 (2020).

37. Lemey, P., Rambaut, A., Welch, J. J. & Suchard, M. A. Phylogeography takes a relaxed random walk in continuous space and time. Mol. Biol. Evol. 27, 1877–1885 (2010).

38. Dellicour, S., Rose, R., Faria, N. R., Lemey, P. & Pybus, O. G. SERAPHIM: studying environmental rasters and phylogenetically informed movements. Bioinformatics 32, 3204–3206 (2016).

39. Rambaut, A. et al. A dynamic nomenclature proposal for SARS-CoV-2 to assist genomic epidemiology. BioRxiv (2020) doi:10.1101/2020.04.17.046086.

40. O’Toole, Å. et al. Assignment of epidemiological lineages in an emerging pandemic using the pangolin tool. Virus Evol. 7, veab064 (2021).

41. Murrell, B. et al. Detecting individual sites subject to episodic diversifying selection. PLoS Genet. 8, e1002764 (2012).

42. Kosakovsky Pond, S. L. & Frost, S. D. W. Not so different after all: a comparison of methods for detecting amino acid sites under selection. Mol. Biol. Evol. 22, 1208–1222 (2005).

43. Shu, Y. & McCauley, J. GISAID: Global initiative on sharing all influenza data - from vision to reality. Euro Surveill. 22, 30494 (2017).

44. Davies, N. G. et al. Estimated transmissibility and impact of SARS-CoV-2 lineage B.1.1.7 in England. Science 372, (2021).

45. Campbell, F. et al. Increased transmissibility and global spread of SARS-CoV-2 variants of concern as at June 2021. Euro Surveill. 26, (2021).

46. Lenth RV. emmeans: Estimated Marginal Means, aka Least-Squares Means,R package version 1.6.1. (2021).

