## Supplementary Material for "Continued Emergence and Evolution of Omicron in South Africa: New BA.4 and BA.5 lineages"

**Extended Data Figures**

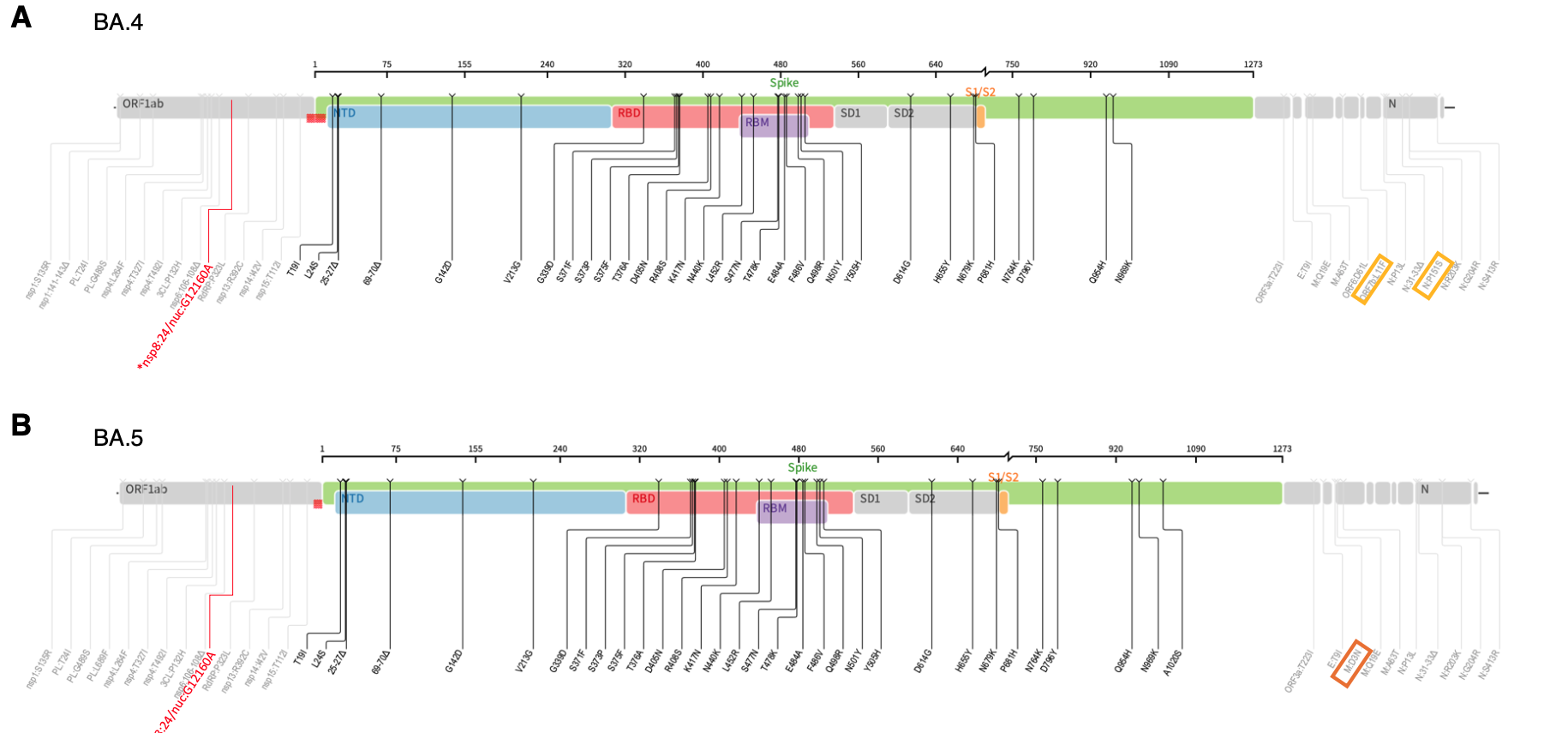

**Extended Data Fig 1:** Whole genome mutations present in BA.4 and BA.5 lineages. Differences in BA.4 and BA.5 are highlighted with a rectangle. The synonymous mutations in nsp8 is indicated in red.

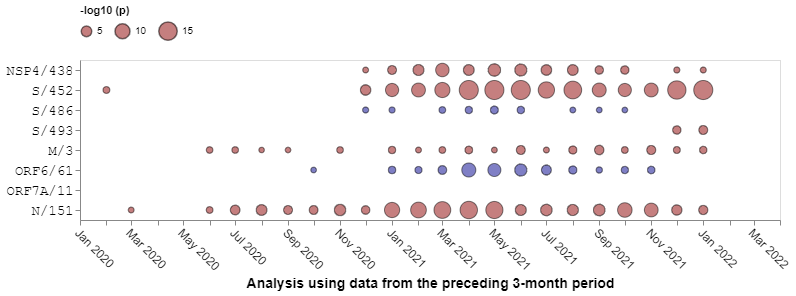

**Extended Data Fig. 2:** Patterns of natural selection between January 2020 and January 2022 at codon sites differentiating BA.4 and BA.5 from BA.2. All SARS-CoV-2 sequences deposited in GISAID were analyzed with each time-point representing an analysis of all sequences sampled during the preceding three months. Red dots indicate evidence at positive selection and blue spots indicate evidence of negative selection. The sizes of the dots indicate degrees of statistical support for selection signals. Only sequences deposited in GISAID prior to the discovery of BA.4 and BA.5 are considered here.

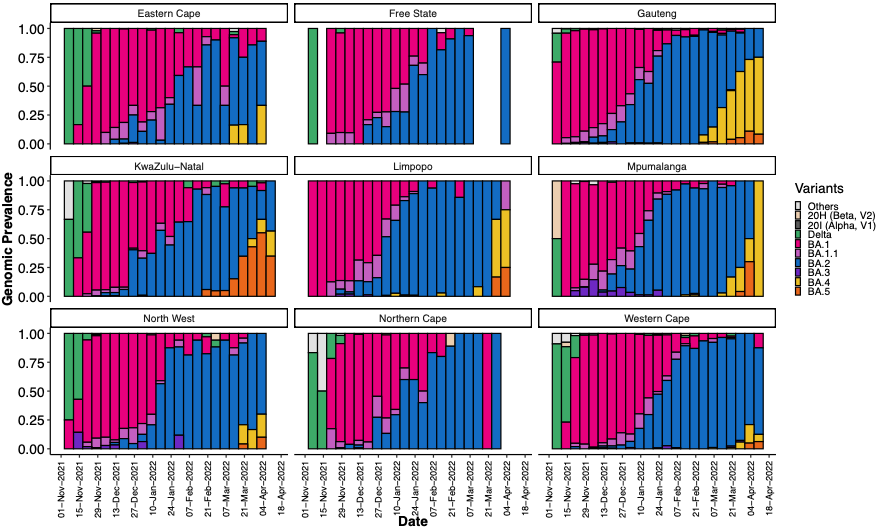

**Extended Data Fig. 3:** Progression of the weekly genomic prevalence of various variants and lineages in the nine provinces of South Africa from November 2021 to April 2022.

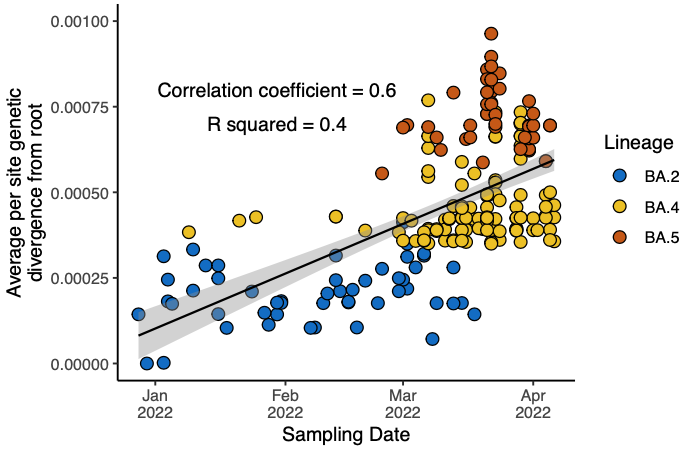

**Extended Data Fig. 4: Molecular clock signal of the dataset of BA.2, BA.4 and BA.5 lineages used in the Bayesian analysis.** Root-to-tip regression obtained from TempEst analysis for the sampled cluster of BA.2, BA.4 and BA.5, showing a relatively strong clock-like behaviour (correlation coefficient = 0.6, R2 = 0.4) The regression line is shown with error buffers (shaded area) that represent 90% confidence intervals.

**Extended Data Table 1:** S-gene target status (TaqPath™ COVID-19 qPCR assay) for 198 samples sequenced by KRISP laboratory

| Omicron lineage | S-gene target failure | S-gene target positive |
| --- | --- | --- |
| BA.1 | 9 | 0 |
| BA.2 | 2* | 120 |
| BA.3 | 0 | 1 |
| BA.4 | 26 | 0 |
| BA.5 | 40 | 0 |
| Total | 77 | 121 |

*One BA.2 sequence had the 69/70del, while the other BA.2 sequence had large gaps in coverage of the spike gene region.
